# “Wow, this could be massive”: Donor-Conceived Families Experience with Unexpected Recontact for Risk of Inherited Cardiac Conditions

**DOI:** 10.64898/2026.11.11.26367578

**Authors:** Lucas A. Mitchell, Amy Ruscigno, Sarah Mulhern, Laura Yeates, Ivan Macciocca

## Abstract

**Introduction:** Recontacting families post-gamete donation, after a genetic diagnosis is made in the donor, can create complex challenges for healthcare providers and families. However, there is little literature that explores the family experience. This study aimed to explore the experiences of parents and donor-conceived young people navigating the recontact process and undergoing predictive genetic testing for an inherited cardiac condition.

**Methods:** Semi-structured interviews were conducted to explore participants’ experiences in unexpectedly being recontacted and undergoing predictive genetic testing for an inherited cardiac condition. Interviews were recorded and transcribed verbatim. A reflexive thematic analysis was performed.

**Results:** Thirteen participants agreed to be interviewed, nine were parents and four were donor conceived young people. Four overarching themes were identified: (1) Out of the Blue, (2) A ripple effect on the family, (3) Calm after the storm, (4) Beneath the Ripples: the psychological impact.

**Discussion:** Interviews provided insights into fertility services recontact and the predictive genetic testing process. From the themes, we conceptualize a ripple effect model. Findings highlight how cardiac genetics and fertility services are evolving, and the critical role of genetic counselors. The recontact process affects the entire family. However, most participants adapted and moved forward from this event.

## Introduction

With the success of the first *in vitro* fertilization (IVF) in the 1970s,^1^ assistive reproductive technologies (ART) have significantly advanced the field of reproductive medicine.^2^ In the last decade the incidence of births conceived through ART has risen in Australia by 55%.^3^ The development of egg vitrification technology has further revolutionised this by giving individuals greater control over the timing of parenthood, whether due to delays for education, career development, infertility, or health issues.^4^ Broader societal acceptance of diverse family structures, including LGBTQIA+ couples and single parents, has also accelerated the demand for gamete donation.^5^

Despite the growing demand of ARTs and gamete donation, several challenges and considerations remain. The success rates for procedures, such as IVF, remain at approximately 30%.^6^ Utilizing ARTs is often characterized with time commitments and financial and emotional burdens for patients.^7^ Ethical debates also continue around gamete ownership, as well as donor identity and the right to initiate contact post-donation.^8^ This is particularly relevant for future recontact between donors and families if clinically significant medical findings are identified.

Cases have been documented where donors, many years post donation, found out they carried a pathogenic variant for a cardiac condition, putting their biological children at 50% risk.^9^

ART laws are governed by state legislation within Australia with support from the National Health and Medical Research Council (NHMRC), which enforce altruistic, non-anonymous gamete donation and place limits on the number of donations from a donor.^10^ Mandatory counseling and informed consent are required for both donors and parents prior to conception. In Victoria, the state legislation enables donor-conceived individuals to access donor information from a centralized state register once they reach 18 years of age. Under exceptional circumstances, ART providers and fertility services may recontact and disclose donor’s medical information to the donor-conceived individual or their family. This disclosure is permissible if a medical doctor determines it is necessary to save a person’s life, to warn them of a genetic or hereditary condition that could be harmful.^11^

Inherited cardiac conditions (ICCs) are a heterogeneous group of disorders with a genetic etiology, encompassing cardiomyopathies and inherited arrhythmias.^12^ Symptoms can range from palpitations and breathlessness to the most severe complications of heart failure and sudden cardiac death. Clinical heterogeneity is common in gene positive individuals within a family.^13^ The identification of a pathogenic / likely pathogenic variant for an ICC can enable the tailoring of clinical management for individuals, and provides at-risk biological relatives the opportunity to undergo predictive testing and clarify their own genetic risk.^14^ Preventive interventions include medications, lifestyle adjustments, implantable cardioverter defibrillators or heart transplantant.^15^ Recent literature identified predictive testing for ICCs profoundly impacts young people’s family relationships and daily self-perception, highlighting a vital need for developmentally appropriate support.^16^

### Rational & Aim

There is little research on donor recontact and predictive genetic testing for at-risk biological children. Notifying donor-conceived families about potential life-threatening cardiac risks presents unique ethical challenges for both families and healthcare providers. This study aims to investigate the experiences of parents and donor-conceived young people navigating the recontact process and undergoing predictive genetic testing.

## Methods

### Design

This qualitative study utilized semi-structured interviews to investigate the experiences of donor-conceived families regarding recontact for predictive genetic testing for an ICC, following a positive genetic result identified in the donor years after conception. Ethics approval was obtained from the Royal Children’s Hospital, Parkville, Victoria, Australia (HREC number: 38299).

### Participants

Participants included donor-conceived young people aged 10 or over (conceived via gamete donation in Victoria, Australia), and parents or legal guardians of a donor-conceived child.

Participants needed to have undertaken the predictive testing process for an ICC and received their result between 1 January 2009 and at least four months before being invited to this study. Participants were required to be proficient in English (see Supplementary File 1).

Participants were identified via clinics at the Victoria Clinical Genetics Service, The Royal Children’s Hospital, Parkville, Victoria, Australia. Eligible participants were invited to join the study via phone call, if interested, they were emailed an invitation pack to join the study. This invitation pack included information about the study and a consent form. Participants returned the forms and consented themselves, or on their child’s behalf. Following consent, the research team scheduled the participant’s interview. If there was no response from prospective participants, the research team followed up two weeks later with three attempts of contact. No response after three attempts was taken as a passive decline (See Figure 1).

**Figure 1.**
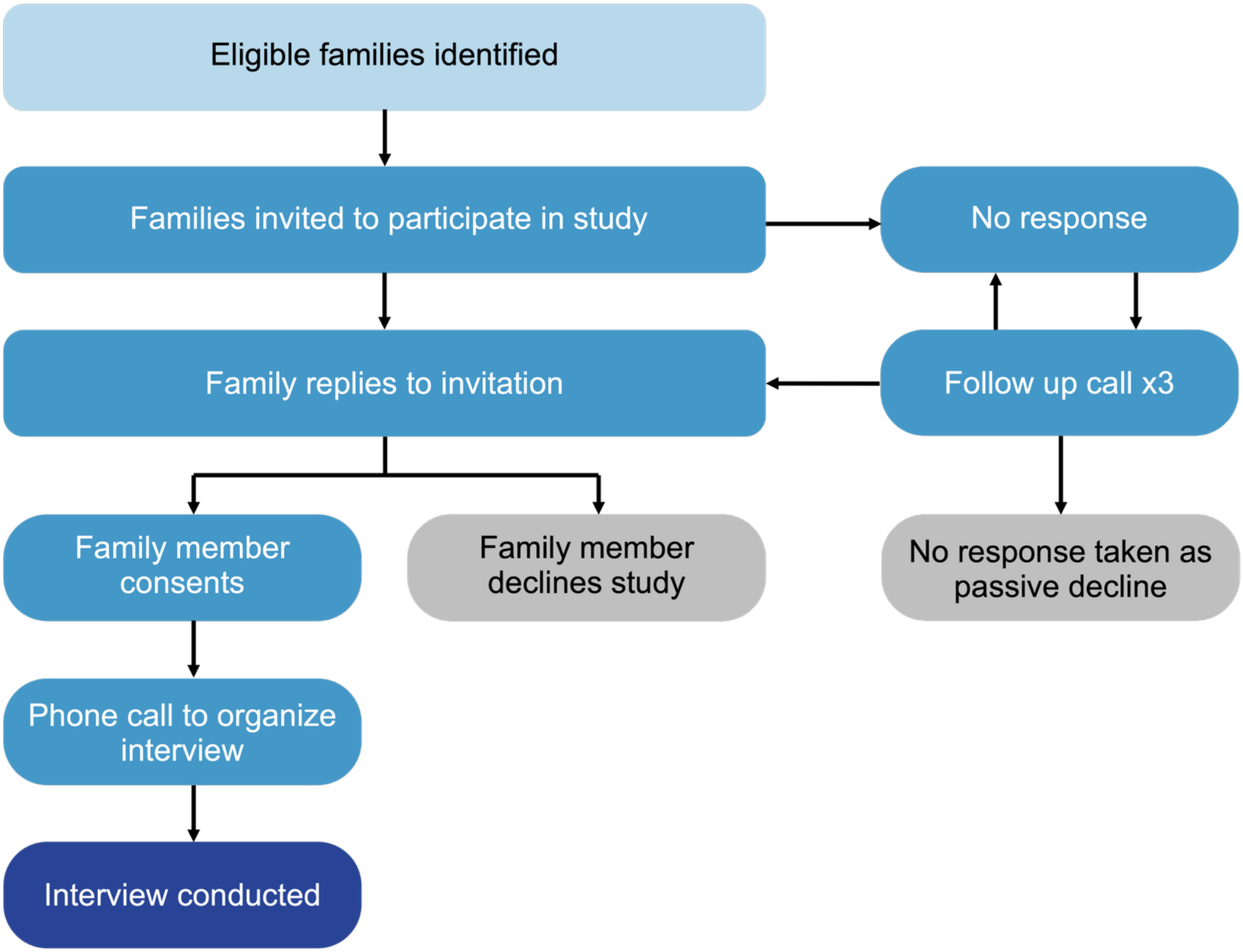
Summary of participant recruitment and interview process

### Data collection

Semi-structured interviews via telephone and video conference software were conducted by AR. Demographic information was collected then followed by a series of open-ended questions regarding participant’s experiences of recontact and undergoing predictive genetic testing for ICCs. Questions focused on the communication of genetic information from healthcare professionals and explored the support offered to participants during and after genetic testing.

The interview guide can be found in Supplementary File 2. Interviews were recorded and transcribed verbatim. Transcripts were checked for accuracy and deidentified.

### Data Analysis

De-identified interview transcripts were uploaded to NVivo 15 software for coding. A reflexive thematic approach was conducted following the six stages outlined by Braun & Clarke.^17^ All interviews were analyzed using an iterative thematic approach. Analysis was conducted by LM, with regular collaborative meetings involving the research team to review and discuss developing codes and themes.

### Reflexivity Statement

The research team consists of genetic counselors from clinical, research and academic institutions. LM is a research genetic counselor and PhD candidate in rare diseases and genomics, with experience in qualitative research and returning genetic research findings to participants. LM has no clinical relationship with any of the participants. AR conducted the interviews and had no prior clinical relationship with participants. The broader research team includes certified genetic counselors (LY, IM, SM) with substantial clinical expertise in cardiovascular genetics, specifically supporting families, including young people. The research team is non-disabled, cisgender women and men of European background residing in Victoria and New South Wales, Australia. To mitigate individual biases, LM continuously refined the analysis through regular supervision and team meetings to ensure interpretation aligned with participants’ raw data.

## Results

### Participants

Twenty-four participants from 13 families were identified as eligible for this study and invited. Nine families consented, while two families declined and two families were lost to follow-up. Within the nine families, nine participants were parents and four were young people. This cohort was entirely of European ancestry (100%) and predominantly female (n=11, 85%). Participants underwent genetic testing for cardiomyopathies (n=7, 54%) and arrhythmias (n=6, 46%), however the majority tested negative (n=10, 77%) for the familial variant causing an ICC. At result disclosure, 55% (n=6) of young people were age 10 or below, and at the time of the interview three of the four young people were older than 10 years of age (n=3, 75%). For most, genetic testing was within the last 1-5 years (n=8, 89%). The initial recontact method from the fertility services varied with five participants contacted by telephone and four received a letter. See Table 1 for summary of characteristics.

**Table 1.**
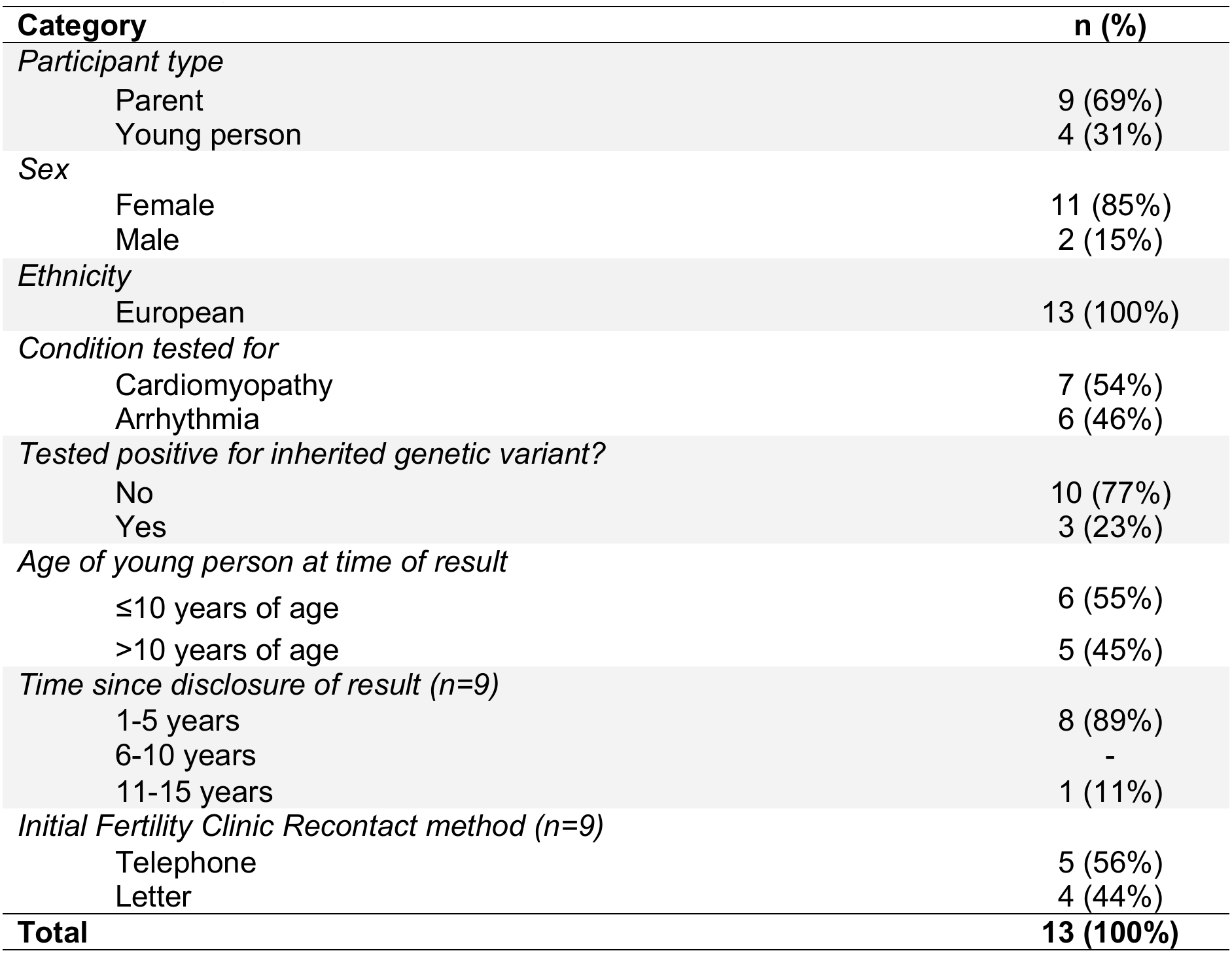
Participant characteristics.

| **Category** | **n (%)** |
|---|---|
| *Participant type* |  |
| Parent | 9 (69%) |
| Young person | 4 (31%) |
| *Sex* |  |
| Female | 11 (85%) |
| Male | 2 (15%) |
| *Ethnicity* |  |
| European | 13 (100%) |
| *Condition tested for* |  |
| Cardiomyopathy | 7 (54%) |
| Arrhythmia | 6 (46%) |
| *Tested positive for inherited genetic variant?* |  |
| No | 10 (77%) |
| Yes | 3 (23%) |
| *Age of young person at time of result* |  |
| ≤10 years of age | 6 (55%) |
| >10 years of age | 5 (45%) |
| *Time since disclosure of result (n=9)* |  |
| 1-5 years | 8 (89%) |
| 6-10 years | - |
| 11-15 years | 1 (11%) |
| *Initial Fertility Clinic Recontact method (n=9)* |  |
| Telephone | 5 (56%) |
| Letter | 4 (44%) |
| **Total** | **13 (100%)** |

### Reflexive Thematic Analysis

From the interviews four themes were identified: (1) Out of the Blue, (2) A ripple effect on the family, (3) Calm after the storm, (4) Beneath the Ripples: the psychological impact. These four themes highlight the different stages of contact and turbulence participants felt throughout this process. See figure 2.

**Figure 2.**
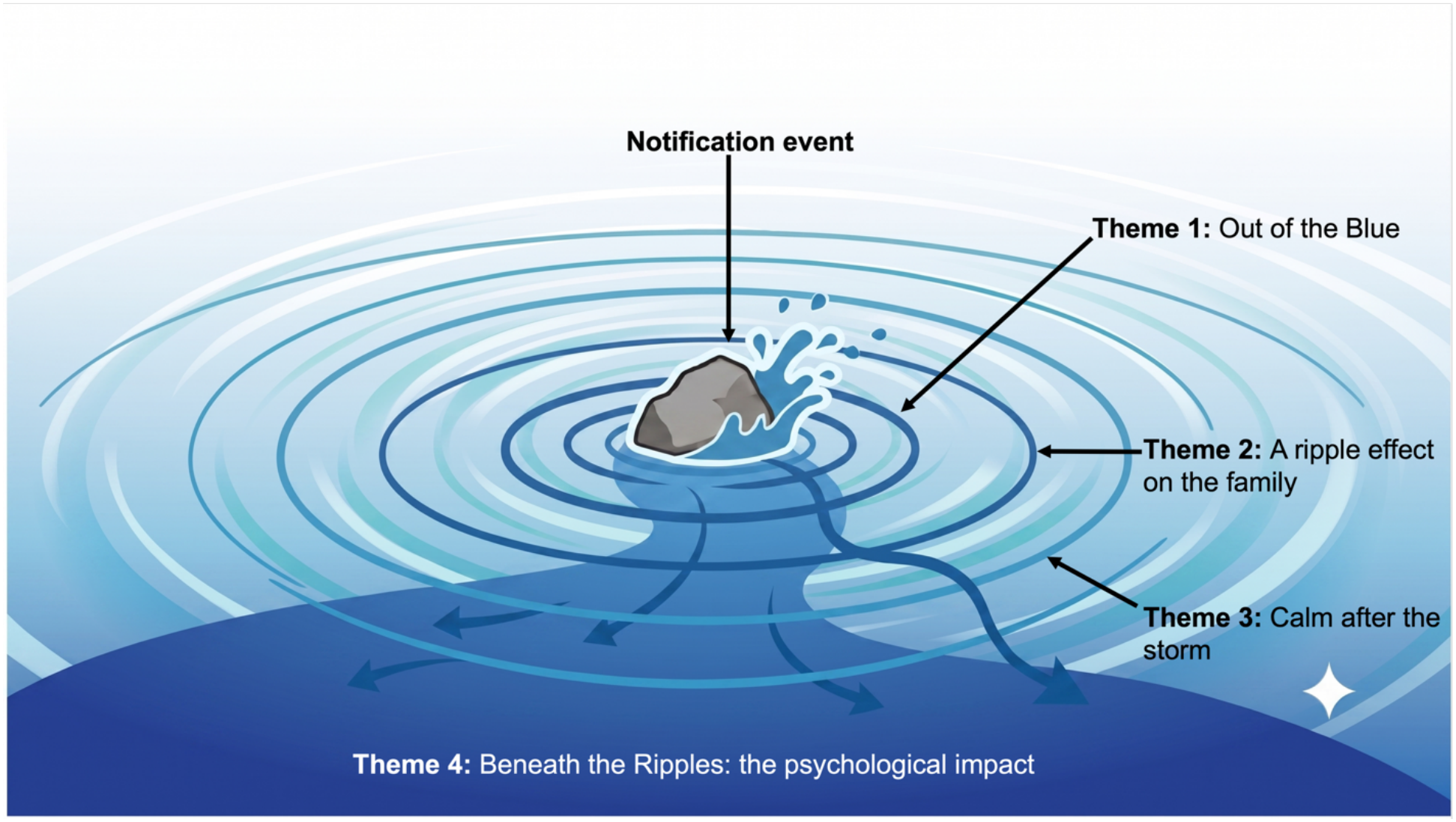
Conceptual ripple model illustrating the identified themes following recontact and predictive genetic testing. Theme 1 (Out of the blue): following immediate notification event (inner ripple). Theme 2 (A ripple effect): Ongoing family adaptation and dynamics (middle ripple). Theme 3 (Calm after the storm): Passing of time and return to normalcy (subsiding outer ripple). Theme 4 (Beneath the Ripples: the psychological impact) the continuous psychological response throughout.

**Figure 3.**
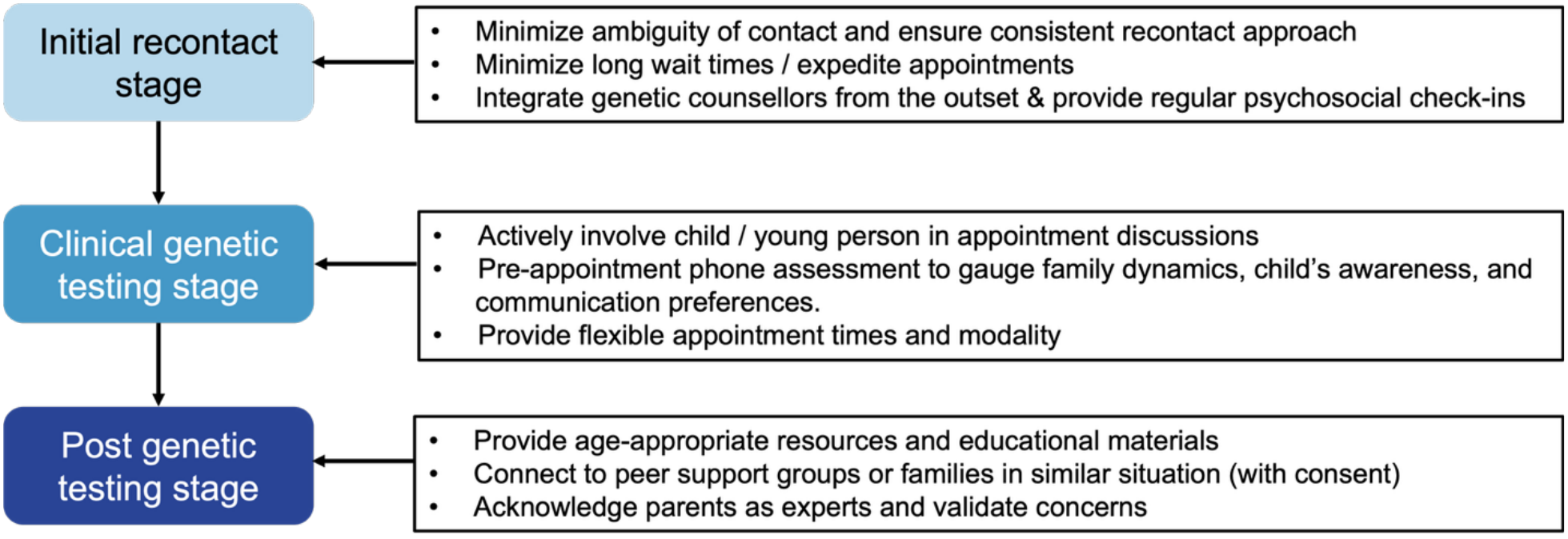
Summary of practice implications for clinicians

#### Theme 1: Out of the Blue

Participants described the initial contact from the IVF service and their method of contact. In many cases participants received a notification letter regarding a potential genetic risk to their child as a sudden and profound shock. This recontact was frequently characterized as something that appeared “out of the blue” as many parents believed their involvement with fertility services had concluded. Therefore, they did not anticipate further communication, or consider the possibility of an ICC.

Upon receiving a registered letter, parents were directed to contact the fertility service for further details. Responses to this initial outreach varied, some families were caught off guard while others remained unconcerned:

> *“I don’t suffer from anxiety or anything like that, but it did make me think, wow, this could be massive.”*
>
> - Parent 8
>
> *“ … I wasn’t too concerned either at the time, I thought OK we’ll just do whatever testing needs to be done.”*
>
> *-* Parent 4

Other parents felt differently and criticized the correspondence as being overly ambiguous and failing to provide sufficient context. This ambiguity heightened uncertainty, leaving parents frustrated when fertility staff could not address specific concerns or were unreachable during office hours. Consequently, families were referred to clinical genetics teams and genetic counselors to navigate the process.

> *“I was just so angry because there was no one to talk to and it was a registered letter so they knew when it would be delivered and I just thought that that was so poorly handled”*

#### *-* Parent 9

Many participants were unaware fertility services could recontact them with future medical updates or had no knowledge of this happening to other families. Verifying and trusting communications from fertility services also proved challenging for some; one parent noted that clinic emails were frequently filtered into junk folders and ignored calls from private numbers due to the prevalence of phone scams, which delayed action. This sense of being caught off guard extended beyond the recontact process itself and to the medical conditions involved, as many parents were completely unfamiliar with the cardiac condition.

> *“So I got a letter, didn’t understand it… actually I did find out beforehand, it’s like, you know, you search on the internet what it is, life expectancy is this or whatever … I guess with the uncertainty, everything, it was really stressful”*
>
> - Parent 5

While some had misconceptions that comprehensive genetic screening was a standard clinical process at the time of donation, or were entirely unsure of the screening process.

> *“No, I don’t actually know… how it is screened, what they screen for. I don’t know any of that sort of stuff.”*
>
> - Parent 2

#### Theme 2: A ripple effect on the family

The experience of being recontacted by fertility services and the subsequent genetic testing was described by both parents and young people as a disruption to family equilibrium. Following the initial notification, participants reflected on how the news evoked complex emotions regarding their donor. Parents, in particular, thought about the logistics of disclosing this risk information to their children or the most appropriate individual.

Being recontacted and undergoing predictive genetic testing for ICCs resurfaced past discussions and feelings towards donor status and identity. While many young people were already aware they were donor conceived, as their parents had clarified the role and relationship of their donor, this was not something they thought about actively.

> *“ I knew about the donor, how I was conceived through donor sperm through IVF… but it wasn’t something that was really front of mind until all of this came up.”*
>
> - Young person 1

Across the fertility services, genetics and cardiology, many parents chose to act as intermediaries, controlling the flow of information to their children. Rather than concealing the truth, parents described delaying disclosure to verify details from the notification. Their priority was to consult with genetic professionals first to ensure they had accurate information before initiating a conversation with their child. Parents also carefully considered the logistics of this disclosure, reflecting on the most suitable timing and environment, such as at home or after school, to share this notification. The existing transparency within some families facilitated a smoother communication process. For these young people, their donor status was already a normalized aspect of their identity, making the notification of a potential ICC a more routine matter. Conversely, parents of younger children or those who had not yet disclosed donor status faced greater challenges in navigating conversations about predictive genetic testing and cardiac screening.

> *“I would have told my husband that night, but I would have waited until the kids went to bed and then we told them separately, and we just explained it really basic.”*
>
> - Parent 7, parent of donor-conceived children

Broader communication and family dynamics was another element of this theme, with parents, and young people being selective in who they shared this risk information with. Some parents described disclosing the situation to their own parents, siblings, or close friends for support during their child’s predictive testing. Meanwhile some young people chose total nondisclosure to their school peers, primarily to avoid ‘opening the can of worms’ in explaining not just testing for an ICC but also their donor-conceived status to their friends.

> *“The only thing I didn’t like was when I went back to school and everyone was asking me what happened and I didn’t want to tell them, so I was like, I was sick and they’re like, OK.”*
>
> - Young Person 4

#### Theme 3: Calm after the storm

Participants discussed their interactions with healthcare professionals and their recollections of clinical appointments. Many participants could not recall specific technical details, however they retained the gist of the conversations. Parents and young people described a desire to move forward rather than dwelling on their genetic result, with some even forgetting the experience entirely. Parents of children with a positive result initially expressed disappointment that their child had a genetic variant for an ICC. However, they quickly focused on gratitude for early intervention and the donor’s proactive approach.

> *“Definitely sad and disappointed that they’ve got the gene but thankful that we found out about it, you know, it could have ended in disaster.”*
>
> - Parent 1

Participants spoke about their engagement with healthcare providers. Many parents appreciated pediatricians and cardiologists for their reassuring communication and ability to ease concerns around their child’s cardiac health. However, a small minority reported negative experiences with specialist follow-up care, describing condescending experiences or having parental concerns dismissed. In clinical genetics, genetic counselors were highly regarded. During periods of uncertainty, the genetic counselor provided clear communication, coordinated next steps, and addressed concerns following the fertility services notifications. Parents felt reassured by the genetic counselors’ expertise and ability to actively engage children in appointments.

> *“I remember liking [genetic counselor] and being reassured by [genetic counselor] and, you know, feeling like [genetic counselor] was giving us all the information that we needed … all of my dealings with [genetic counselor] were good.”*
>
> - Young person 1

Both parents and young people identified logistical hurdles during the testing process, offering recommendations to refine future care. A key point of discussion was the clinical genetic appointments. Many participants valued in-person hospital appointments, noting that face-to-face interaction provided clarity and helped resolve uncertainties. However, others found traveling to the hospital challenging, particularly those residing in regional or remote areas.

Some young people also found the hospital environment intimidating and expressed a preference for having their parent present for support:

> *“I probably would have rather have mom there because I don’t like hospitals, there’s always something beeping and then there’s always someone talking”*
>
> - Young Person 4

Some parents expressed dissatisfaction with waiting for a formal results appointment, and would have preferred receiving the result via telephone to avoid delays. Participants emphasized the importance of a genetic counselor or a clinical genetics team to ensure clear communication and robust support throughout the testing process. Some even suggested the initial notification from the fertility service should ideally be handled directly by a genetic counselor. Participants believed this approach would have provided more comprehensive context from the outset, rather than a referral to another healthcare professional.

> *“I think [genetic counselor]’s phone call first might have been better … Like [genetic counselor] was able to answer all of those questions for me.”*
>
> - Parent 2

Participants identified specific areas where additional support would have been beneficial. These included more frequent check-ins and guidance during the initial contact phase and the period leading up to the first clinical genetics appointment, as these stages were characterized by high anxiety. Participants expressed a desire for connection to other families, and highlighted that access to specialized support groups would have been valuable. Additionally, access to online and take-home resources was also recommended.

#### Theme 4: Beneath the Ripples: the psychological impact

Throughout this whole process, parents and young people reflected on their emotional reactions and had varying ongoing psychological impact. Anxiety and stress were common sentiments reported across the entire process, beginning with the unexpected notification and continuing through the testing and waiting periods for results. Some participants spoke of externally appearing fine, however underneath they were stressed during this period along with their wider family:

> *“On the outside I was fine, you know, going through the motions. But on the inside, I was like, oh my gosh. And not just me, like Mum and my brother and you know, we were all stressed”*
>
> - Parent 1

In response, participants described various coping mechanisms. While some engaged in active information-seeking to gain clarity, others intentionally avoided reading excessively online about ICCs to mitigate anxiety and maintain a balanced perspective. Parents frequently described a protective response or effort to maintain a brave face for their child. Conversely, some young people wanted to avoid worrying their parents by attempting to appear brave during clinical appointments and testing, which was simultaneously observed by parents.

> *“[Son] wasn’t too concerned about having all the tests done. Yeah, he knew that I was very worried and stressed about it and I think he hid a lot of his emotions from me at the time.”*
>
> - Parent 3

## Discussion

This study investigated the parents and donor-conceived young people’s experiences in being recontacted by fertility services and undergoing subsequent predictive genetic testing for an ICC. From our analysis we conceptualized a water ripple metaphor that conveys this experience. This metaphor captures the immediate shock of unexpected recontact (Theme 1: Out of the Blue) and its subsequent ripple effect on parent-child relationships and family communication (Theme 2: A ripple effect on the family). Following predictive testing and result disclosure, families eventually experienced a return to normalcy (Theme 3: Calm after the storm), while highlighting their psychological response throughout this process (Theme 4: Beneath the Ripples: the psychological impact).

Many participants expressed shock and confusion regarding the unexpected, ‘out of the blue’ contact from fertility services and reported not anticipating further communication. Meanwhile some had assumed genetic testing on gametes had already been performed at conception. This aligns with a 2024 qualitative genetic breast cancer study, which also highlighted this ‘out of the blue’ response to recontact.^18^ A 2025 mixed-methods preconception carrier screening study also found parents were unfamiliar with screening tests. Some believed carrier screening could identify a reason for their fertility problems or increase their chance of a pregnancy.^19^ These findings highlight a broader lack of societal ART literacy, which may have partly contributed to parents’ misconceptions and response to the initial contact.

Our study highlighted the importance of a genetic counselor throughout this process. Many participants spoke highly of genetic counselors and preferred to have spoken to them from the outset, rather than the fertility service. Genetic counselors are ideally placed for these conversations, with training and expertise in counseling, adaptation, emotional support, as well as advanced genetic knowledge to support families.^20^ Research has shown that continuity of care from healthcare providers also leads to better patient satisfaction and reduced uncertainty for patients.^21^ Having the same healthcare professional from the recontact through to testing could have eased parental anxieties from the outset.

In our sample only a small number of parents had not disclosed donor conception to their children prior to recontact, however this did not significantly impact overall coping. A previous meta-analysis investigating parental disclosure of ART conception to their children (aged 1-22), found 77% of parents had not disclosed yet or had plans to disclose to their child.^22^ Earlier awareness of donor status has been associated with better psychological well-being within the child,^23^ and less negative emotions reported.^24^ Children learning of their donor status after age 18 could not agree on an ideal age to find out, but wished they had known sooner.^25^ This literature may partly explain why most of our young participants adjusted well to this information over both the short and long term, as most families were already open with use of gamete donation.

The impact of the notification and process of testing on the family unit can possibly be understood through family systems theory.^26^ This framework highlights the interconnected dynamics of families and that individuals do not operate in isolation. In particular, the triangulation of relationships, parental projections and communication can shape individual responses.^26^ Further foundational work by Rolland’s (1987) Family Illness Paradigm also highlights “mastery of control” as a key element here in how families navigate uncertainty.^27^ In this paradigm, we can see how parents may act as intermediaries to communicate risk information to their child. However, many of our participants also appreciated when genetic counselors spoke directly to and included the young person in appointments.

Although the testing process caused a potential disruption within the family, our findings indicate that families’ anxieties and concerns gradually subsided over time. Many participants retained the general clinical messages or “gist” of their consultations, but did not recall specific technical details. This adaptation aligns with broader literature on adjustment to post predictive testing, as well as, genetic findings from research, which has demonstrated participants adjust to this information over time with minimal long-term negative impact.^18,28–31^

## Limitations

The cohort of this study was small and entirely of European ancestry (100%), predominantly female (85%), and resided in Victoria, Australia, meaning males, and culturally diverse families are underrepresented. It is also worth noting only sperm-donor conceived families were interviewed. Given the different Australian state laws around ART and state-based funded healthcare systems, families in other states of Australia or other countries may have different experiences. Findings in our study heavily reflect parental perspectives (69%) compared to young people, and most families had a negative genetic result (77%). This overrepresentation of negative results potentially influences our findings, as families managing a positive diagnosis may have different long-term care experiences. Finally, the cross-sectional snapshot does not provide long-term follow-up, and the retrospective nature of interviews could lead to potential recall bias regarding past events and the emotional impact of recontact and testing.

## Practice Implications

Several key implications were identified. Firstly, participants highlighted a preference speaking to a genetic counselor from the outset, which could better assist families in clarifying genetic risk information and processing uncertainty. Participants also wanted more support during the initial phase when they had spoken to fertility services but were still waiting for their genetics appointment. More psychological support and contact from a genetic counselor should be offered during this waiting time. These interviews also underscored the importance of a pre-appointment phone call. Participants expressed high regard for the genetic counselor who reached out beforehand to understand the unique family dynamics and the child’s awareness of their donor status. This proactive contact successfully informed the discussion and tailored the approaches in the clinical appointment. Participants appreciated when clinicians took the time to recognize the unique family dynamics, structures and communication preferences among parents and children. Families discussed the need for specialized, take-home or online resources to help them explain complex genetic information and gamete donation to their children. Families also desired for more connection with people who are in similar situations. However, given this is a small cohort of people affected, future research should investigate safe opt-in frameworks to facilitate peer connection without compromising families’ privacy.

## Conclusion

This study provides in-depth insights from parents and donor-conceived young people regarding their experiences of recontact by fertility services and subsequent predictive genetic testing for an ICC. Our findings were conceptualized in a ripple effect model, highlighting the impact on the family unit and that over time most participants adapted and moved forward. Importantly, the psychological responses show the unique and varied experiences of participants throughout this process. Genetic counselors were seen as key healthcare professionals in this process, however, were underutilized in the initial contact period. The active inclusion of young people in clinical appointments, more take-home resources, and peer support groups was highly desired.

## Data Availability Statement

Anonymized redacted transcripts can be made available to the authors upon request and with appropriate human research ethics approval.

## Acknowledgments

We would like to thank the parents and young people for their participation. This work was conducted in part to fulfill a requirement of a Doctor of Philosophy for LM who is also supported by a National Health and Medical Research Council (NHMRC) PhD Scholarship (#2039978). This work was also conducted in part to fulfill a requirement for the degree of Master of Genetic Counselling for AR.

## Author Contribution Statement

Conceptualization: IM, LY, SM; Data curation AR; Formal analysis: LM; Methodology: IM, LY, SM; Project administration: AR, LM; Supervision: IM, LY; Visualization: LM; Writing-original draft: LM; Writing-review & editing: LM, AR, SM, IM, LY

## Ethical Approval

Ethics approval was obtained from the Royal Children’s Hospital, Parkville, Victoria, Australia (HREC number: 38299). This study was conducted with strict accordance to ethical conduct in human research. All adult participants provided written informed consent. For young people under the age of 18, parent/guardian written consent was obtained.

## AI Statement

During the preparation of this work the authors used an institutional license of Google Gemini to assist with spelling, grammar and generation of figure 2. After using AI, the authors reviewed and edited the content as needed and take full responsibility for the content within this publication.

