## Supplementary information for "“Wow, this could be massive”: Donor-Conceived Families Experience with Unexpected Recontact for Risk of Inherited Cardiac Conditions"

**Supplementary Table 1**. Participant inclusion and exclusion criteria

| Inclusion Criteria | Exclusion Criteria |
|---|---|
| • Used sperm donation in Victoria, Australia<br>• Donor-conceived child underwent PT for an ICC through VCGS and received their result<br>• After January 1, 2009 and at least 4 months before being approached for recruitment<br>• Fluent in English<br>• Is a parent or legal guardian of a young person who meets the above criteria<br>• Young person was aged 10 or over at time of interview | • Documented medical history of a mental or psychiatric illness within 12 months of recruitment<br>• Had experienced a recent loss of a family member due to an ICC 12 months prior to recruitment |

### Interview Guide

These are sample questions for the benefit of the HREC panel only. They will not be provided to participants prior to the interview.

The interviews are semi-structured and as such will not adhere to a strict interview schedule. Questions may be asked out of order in the interest of conversational flow, and in the course of answering one question a participant may address another. As participants have a range of experiences, not all sample questions listed below are applicable. Therefore, not every participant will be asked every question, and questions not included in the below sample may be asked if relevant and natural. As interviews are completed, this interview guide may be modified to incorporate key issues raised in earlier interviews.

Questions with a bullet point are indicative prompts which may be used to elicit more information regarding the numbered questions, if necessary.

All interviews will address the following:

- Demographics
- Experience of learning about the inherited cardiac condition
- Experience of the process of predictive testing & health professionals
- The implications of testing
- Perceptions of healthcare team and support

| TOPIC | PARENT | YOUNG PERSON<br>NB: Questions will be adapted according to age/developmental stage |
|---|---|---|
| *Demographics* | 1. To start off, could you please tell me a little bit about yourself<br>● Age, children, relationship | 1. Tell me a little bit about yourself<br>● Age, school, siblings |
| *Experience of learning about the inherited cardiac condition* | 2. Can you tell me a little bit about how you learnt that [child] had a chance of inheriting [ARVC/Long QT/ HCM]<br>○ How was that experience for you?<br>○ How do you think that experience was for [child]?<br>○ Did you tell your child?<br>○ If you told your child:<br>■ How was it communicating this information?<br>■ What do you think it was like for them hearing this information?<br>■ Do you feel like this was the most appropriate way to share the information?<br>○ If you didn't tell your child:<br>■ What was the main reason of not telling [child]?<br>○ What was your initial reaction/how did you feel?<br>○ What was [child's] initial reaction from your perspective?<br>○ Did you share this information with anyone else?<br><br>*If family are from donor conceived cohort:*<br>3. Were you informed about the possibility of the sperm donor having a medical condition after they donated? | 2. Can you tell me a little bit about how you learnt that you had a chance of having [ARVC/Long QT/HCM]<br>● How was that experience for you?<br>● Who told you?<br>● How did they tell you?<br>● Did you feel like this was the most appropriate way to be told?<br>○ If not how would you have liked to be told, if at all?<br>● What was your initial reaction/how did you feel? (sad, surprised, angry etc)<br>● Did you share this information with anyone? |

| | | |
|---|---|---|
| *Experience of process of predictive testing & health professionals* | 4. Can you tell me a little bit about what happened after you learnt there was a chance [child] inherited [ARVC/Long QT/HCM?].<br>● What did the doctors/health professionals/genetic counsellors tell you and/or [child] about the heart condition in your family?<br>○ How did that make you feel?<br>○ How do you think that made [child] feel?<br>● Do you think [child] remembers having a genetic test?<br>○ Had [child] had any experience with genetic testing before?<br>○ Who told them the result of the genetic test?<br>○ Where were they?<br>○ How do you think it made them feel hearing the result of the genetic test?<br>○ How did it make you feel hearing the result of the genetic test?<br>● Did you see any doctors/ health professionals / genetic counsellors? If so, how was that?<br>● Did [child] talk to health professionals on their own or with someone?<br>○ Do you think [child] would have wanted that? | 4. Can you tell me a little about what happened after you learnt there was a chance of you inheriting [ARVC/Long QT/ HCM]?<br>● What did the doctors/health professionals/genetic counsellors tell you about the heart condition in your family?<br>○ How did that make you feel?<br>● Do you remember having a genetic test?<br>○ Have you had any experience with genetic testing before?<br>○ Who told you the result of the genetic test?<br>○ Where were you?<br>○ How did it make you feel hearing the result of the genetic test?<br>● Did you see any other doctors/ health professionals / genetic counsellors? If so, how was that?<br>● Did you get to talk to the health professionals on your own without your parents?<br>○ Would you have wanted that? |

<table>
<tr><td>Implications of testing</td><td>5. Have there been any changes for [child] since receiving their genetic test result?<ul><li>Lifestyle changes?</li><li>If yes, how do you think [child] feels about these changes?</li><li>Has this result impacted other family members/ had an impact beyond [child]?</li></ul>If family are from donor conceived cohort:<br>6. Did this experience result in any changes to the relationship you may or may not have had with the donor?<ul><li>If positive for family mutation</li></ul>7. Does [child] continue to see a doctor or health professional?<ul><li>How often?</li></ul></td><td>5. Have there been any changes for you since receiving your genetic test result?<ul><li>Lifestyle changes? (eg. sport, school, social)</li><li>If yes, how do you feel about these changes?</li></ul>If positive for family mutation<br>7. Do you continue to see a doctor or health professional?<ul><li>How often?</li></ul></td></tr>
<tr><td>Perceptions of healthcare team and support</td><td>8. Was any kind of support offered to you and [child] after receiving the genetic test result?<ul><li>Post test counselling?</li><li>Form of follow up from a health professional?</li><li>If family are from donor conceived cohort: Information about the donor/resources to connect with other donor families?</li><li>Resources given to understand more about the condition?</li></ul>9. Who did you go to for support?<ul><li>Do you know anybody else in the same situation as you?</li><li>Have you talked to anyone else who has this genetic change?</li></ul></td><td>8. Was any kind of support offered to you after you received your genetic test result?<ul><li>Post test counselling?</li><li>Form of follow up from a health professional?</li><li>If family are from donor conceived cohort: Information about donor conception/resources to connect with other donor families?</li><li>Resources given to understand more about the condition?</li></ul>9. Who did you go to for support?</td></tr>
</table>

| | | |
|---|---|---|
| | 10. Did you feel like [child] understood what the health professional was saying when they received their genetic result?<br>● Did they feel comfortable to ask questions?<br><br>11. If you could magically create any kind of service or support for families like yours, what would that look like?<br><br>*If positive for family mutation*<br>12. Does [child] receive any ongoing support through the hospital, for example through the cardiology department?<br>● If so, how have you found that?<br><br>If relevant based on child age:<br>13. Have you discussed transitioning to adult care? What do you think about that? / Have you transitioned to adult care? How did that go? | ● Do you know anybody else in the same situation as you?<br>● Have you talked to anyone else who has this genetic change?<br><br>10. Did you feel like you understood what the health professional was saying when you received your genetic result?<br>● Did you feel like you could ask questions?<br><br>11. If you could magically create any kind of service or support for young people like you, what would that look like?<br><br>*If positive for family mutation*<br>12. Do you receive any ongoing support through the hospital, for example through the cardiology department?<br>● If so, how have you found that?<br><br>If relevant based on child age:<br>13. Have you discussed transitioning to adult care? What do you think about that? / Have you transitioned to adult care? How did that go? |
